# Inequities in childhood immunisation coverage associated with socioeconomic, geographic, maternal, child, and place of birth characteristics in Kenya

**DOI:** 10.1101/2021.02.14.21251721

**Authors:** Simon Allan, Ifedayo M. O. Adetifa, Kaja Abbas

## Abstract

**Background:** The global Immunisation Agenda 2030 highlights coverage and equity as a strategic priority goal to reach high equitable immunisation coverage at national levels and in all districts. We estimated inequities in full immunisation coverage associated with socioeconomic, geographic, maternal, child, and place of birth characteristics among children aged 12-23 months in Kenya.

**Methods:** We analysed full immunisation coverage (1-dose BCG, 3-dose DTP-HepB-Hib (diphtheria, tetanus, pertussis, hepatitis B and *Haemophilus influenzae* type B), 3-dose polio, 1-dose measles, and 3-dose pneumococcal vaccines) of 3,943 children aged 12–23 months from the 2014 Kenya Demographic and Health Survey. We disaggregated mean coverage by socioeconomic (household wealth, religion, ethnicity), geographic (place of residence, province), maternal (maternal age at birth, maternal education, maternal marital status, maternal household head status), child (sex of child, birth order), and place of birth characteristics, and estimated inequities in full immunisation coverage using bivariate and multivariate logistic regression.

**Results:** Immunisation coverage ranged from 82% [81–84] for the third dose of polio to 97.4% [96.7–98.2] for the first dose of DTP-HepB-Hib, while full immunisation coverage was 68% [66–71] in 2014. After controlling for other background characteristics through multivariate logistic regression, children of mothers with primary school education or higher have at least 54% higher odds of being fully immunised compared to children of mothers with no education. Children born in clinical settings had 41% higher odds of being fully immunised compared to children born in home settings. Children in the Coast, Western, Central, and Eastern regions had at least 74% higher odds of being fully immunised compared to children in the North Eastern region, while children in urban areas had 26% lower odds of full immunisation compared to children in rural areas. Children in the middle and richer wealth quintile households were 43–57% more likely to have full immunisation coverage compared to children in the poorest wealth quintile households. Children who were sixth born or higher had 37% lower odds of full immunisation compared to first-born children.

**Conclusions:** Children of mothers with no education, born in home settings, in regions with limited health infrastructure, living in poorer households, and of higher birth order are associated with lower rates of full immunisation. Targeted programmes to reach under-immunised children in these subpopulations will lower the inequities in childhood immunisation coverage in Kenya.

## Introduction

The total population in Kenya in 2019 was 43.7 million, with 30.2 million people living in rural areas and 13.5 million people living in urban areas [1]. Vaccines have played a significant role in increasing the life expectancy and reducing the under-five mortality rate in Kenya. The life expectancy at birth was 66.7 years and the under-five mortality rate was 45 per 1,000 live births in 2019 [2]. There has been more than a 50% reduction in under-five mortality rate from 1,869 to 831 deaths per 100,000 from 2000 to 2019 among under-5 year-old children [3].

### Expanded programme on immunisation in Kenya

The World Health Organization (WHO) established the Expanded Programme on Immunisation (EPI) in 1974 to improve access to immunisation services worldwide [4]. Kenya launched its EPI program in 1980 to improve and expand immunisation for six priority diseases - diphtheria, measles, polio, tetanus, tuberculosis, and pertussis, and the number of vaccines has since expanded [5]. New vaccines have been introduced to the routine immunisation programme since 2000. Kenya was the first country to launch the pentavalent vaccine (DTP-HepB-Hib – diphtheria, tetanus, pertussis, hepatitis B and *Haemophilus influenzae* type B) with support from Gavi, the Vaccine Alliance in 2001 [6]. Since then, it has added the second dose of measles in 2013, rotavirus in 2014, and inactivated polio vaccine in 2015 [7]. As a signatory to the Global Vaccine Action Plan, Kenya has committed to fully immunising 90% of all children by 2020, with immunisation coverage of at least 80% in each administrative county [8]. WHO and the United Nations Children’s Fund (UNICEF) estimate that the coverage of three doses of Diphtheria-Pertussis-Tetanus (DPT3) in Kenya – a commonly used metric for immunisation coverage – increased from 82% in 2000 to 92% in 2019 [9].

### Inequities in immunisation coverage

The global Immunisation Agenda 2030 highlights coverage and equity as a strategic priority goal to reach high equitable immunisation coverage at national levels and in all districts, and protect everyone with full immunisation, regardless of location, age, socioeconomic status or gender-related barriers [10]. Reasons related to non-vaccination and under-vaccination of children in low and middle income countries include immunisation systems, family characteristics, parental attitudes and knowledge, and limitations in immunisation-related communication and information [11]. A review conducted in collaboration with WHO attributed under-vaccination with factors related to access to services, health staff attitudes and practices, reliability of services, false contraindications, parents’ practical knowledge of vaccination, fear of side effects, conflicting priorities and parental beliefs [12].

Improvements in national immunisation coverage mask differences in coverage between population sub-groups in Kenya. In 2014, there was a 17.7 percentage point difference in DPT3 immunisation coverage between the highest coverage in Central province and the lowest in North Eastern province [13]. With almost 1.5 million children born each year in Kenya, relatively small proportional differences in immunisation coverage between subgroups translates into large absolute numbers of under-immunised children [14]. Given the disparities in DPT3 immunisation, Gavi has identified Kenya as a priority country for support in achieving high and equitable immunisation coverage, and has invested more than USD 500 million to strengthen routine and campaign immunisation services in Kenya [15,16].

### Study objective

Our study objective is to analyse full immunisation coverage among children aged 12-23 months in Kenya and estimate the inequities in full immunisation coverage associated with socioeconomic, geographic, maternal, child, and place of birth characteristics using data from the 2014 Kenya Demographic and Health Survey. Full immunisation encompasses one dose of Bacillus Calmette– Guérin (BCG), three doses of DTP-HepB-Hib, three doses of polio (excluding the birth dose), one dose of measles, and three doses of pneumococcal vaccines, for which vaccine coverage data is available in the 2014 Kenya DHS. We expect the results to highlight the hidden inequities in immunisation coverage, help to identify underserved subpopulations, and provide evidence for informing health policy and practices to improve immunisation coverage and equity in Kenya.

## Methods

### Survey data

The Demographic and Health Surveys (DHS) Program has collected nationally representative data on population health through more than 400 surveys in over 90 countries [17]. These surveys provide estimates of key indicators that cover population, maternal and child health issues.

We analysed the Kenya DHS 2014 dataset to infer inequities in childhood immunisation coverage associated with socioeconomic, geographic, maternal, child, and place of birth characteristics in Kenya [18]. This is the most recent complete Kenya DHS survey and was conducted from May to October 2014; the 2021 survey is in progress. Sampling was conducted in two stages to achieve representativeness at the national, regional, and county levels. The first stage involved the random sampling of 1,612 enumeration areas from 96,251 areas across Kenya. The second stage involved the random sampling of 25 households from each enumeration area. This resulted in a total sample of 40,300 households.

Survey data regarding children, including their immunisation status, was obtained from interviews with the 31,079 eligible women in the sampled households. Data on the immunisation status of children was collected using written immunisation records and, for children where this was not possible, using verbal reports from mothers.

### Full immunisation coverage

We analysed full immunisation coverage among children aged 12-23 months in Kenya. Full immunisation refers to one dose of BCG, three doses of DTP-HepB-Hib, three doses of polio (excluding the birth dose), one dose of measles, and three doses of pneumococcal vaccines. We did not consider the rotavirus vaccine since it was introduced in Kenya’s routine immunization schedule only in July 2014, after surveying had begun.

The pentavalent vaccine uptake is recorded for the combination vaccine (and not for individual antigens), and thereby DTP coverage is assumed from pentavalent coverage. While the 2014 Kenya DHS data set includes coverage data for first, second, and third doses of DTP, this is based on the coverage of the pentavalent vaccine and thereby used as the measure for the pentavalent vaccine coverage. Also, the coverage metrics for DTP3, HepB3, and Hib3 from the WHO and UNICEF Estimates of National Immunization Coverage (WUENIC) are similar for Kenya in 2014 [19].

### Equity criteria

We used equity criteria based on the WHO’s guidance on priority-setting in health care (WHO GPS-Health) in relation to social groups and a WHO assessment of inequalities in childhood immunisation in ten Gavi priority countries [13,20]. We selected the following explanatory variables: household wealth, religion, ethnicity, place of residence, location/region, maternal characteristics (age at childbirth, education, marital status, position within household), sex of child, birth order, and place of birth.

### Survey analysis

We disaggregated mean coverage by socioeconomic (household wealth, religion, ethnicity), geographic (place of residence, location/region), maternal (age at childbirth, education, marital status, status within household), child (sex of child, birth order), and place of birth. We conducted simple and multivariate logistic regression to assess associations between full immunisation coverage and socioeconomic, geographic, maternal, child, and place of birth characteristics. Simple logistic regression was used to estimate crude odds ratios and multivariate logistic regression was used to estimate adjusted odds ratios. Two variables, sex of the child and maternal age at birth, were selected *a priori* for the multivariate logistic regression based on the findings from previous studies [21–23]. Tests were conducted for collinearity between explanatory variables and collinear variables were removed from the model. Thereby, we estimated adjusted odds ratios using multivariate logistic regression to infer inequities in full immunisation coverage (1-dose BCG, 3-dose DTP-HepB-Hib, 3-dose polio, 1-dose measles, and 3-dose pneumococcal vaccines) in Kenya among children aged 12–23 months associated with socioeconomic (household wealth), geographic (place of residence, province), maternal (maternal age at birth, maternal education, maternal marital status, maternal household head status), child (sex of child, birth order), and place of birth characteristics.

Sampling weights were applied to the survey dataset to adjust for disproportionate sampling and non-response and ensure the sample was representative of the population. The survey analysis was conducted using the Stata statistical software [24] and visualisations were generated using the R statistical software [25]. The analysis code is publicly accessible at https://github.com/vaccine-impact/vaccine_equity_kenya and the 2014 Kenya DHS data set is accessible upon registration on the DHS website at https://www.dhsprogram.com/methodology/survey/survey-display-451.cfm.

### Ethical considerations

This study was approved by the ethics committee (Ref 19139) of the London School of Hygiene & Tropical Medicine. In general for DHS surveys, the survey procedures and questionnaires are approved by the ICF Institutional Review Board (IRB), and the country-specific DHS survey protocols are reviewed by the ICF IRB and an IRB in the host country.

## Results

### Childhood immunisation coverage

Immunisation status data was collected for 3,965 living children aged 12-23 months in the 2014 Kenya DHS Survey. We excluded data for 22 children who had missing or unknown data for at least one vaccine across the full recommended course of routine vaccines. Therefore, we conducted our analysis using data for 3,943 children for which immunisation status data were available for all vaccines included in full immunisation (1-dose BCG, 3-dose DTP-HepB-Hib, 3-dose polio, 1-dose measles, and 3-dose pneumococcal vaccines). The proportions of male and female children were similar at 52% and 48% respectively.

Table 1 presents the coverage for individual doses of the BCG, DTP-HepB-Hib, polio, measles, and pneumococcal vaccines. Full immunisation coverage was 68% [66–71] in 2014. Single immunisation coverage ranged from 82% [81–84] for the third dose of polio to 97.4% [96.7–98.2] for the first dose of DTP-HepB-Hib. Figure 1 shows full immunisation coverage in the eight regions (defunct provinces) of Kenya. Full immunisation coverage ranged from 42% [36–49] in the North Eastern region to 78% [72–83] in the Central and Eastern regions.

**Table 1:**
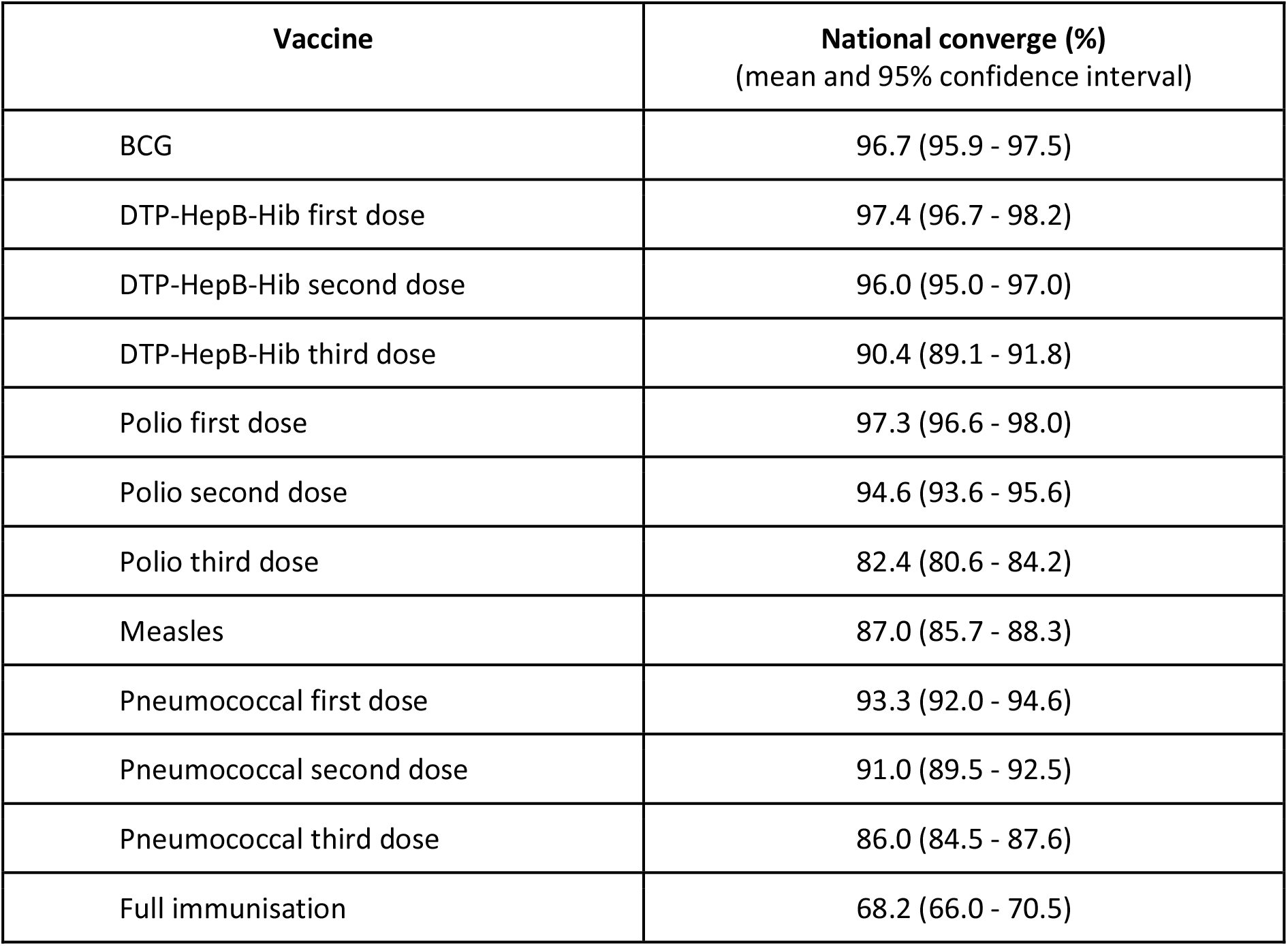
Immunisation coverage in Kenya at the national level. Immunisation coverage (mean coverage and 95% confidence intervals) in Kenya among children aged 12-23 months based on a nationally representative sample of 3,943 children. Full immunisation includes 1-dose BCG, 3-dose DTP-HepB-Hib, 3-dose polio, 1-dose measles, and 3-dose pneumococcal vaccines.

**Figure 1:**
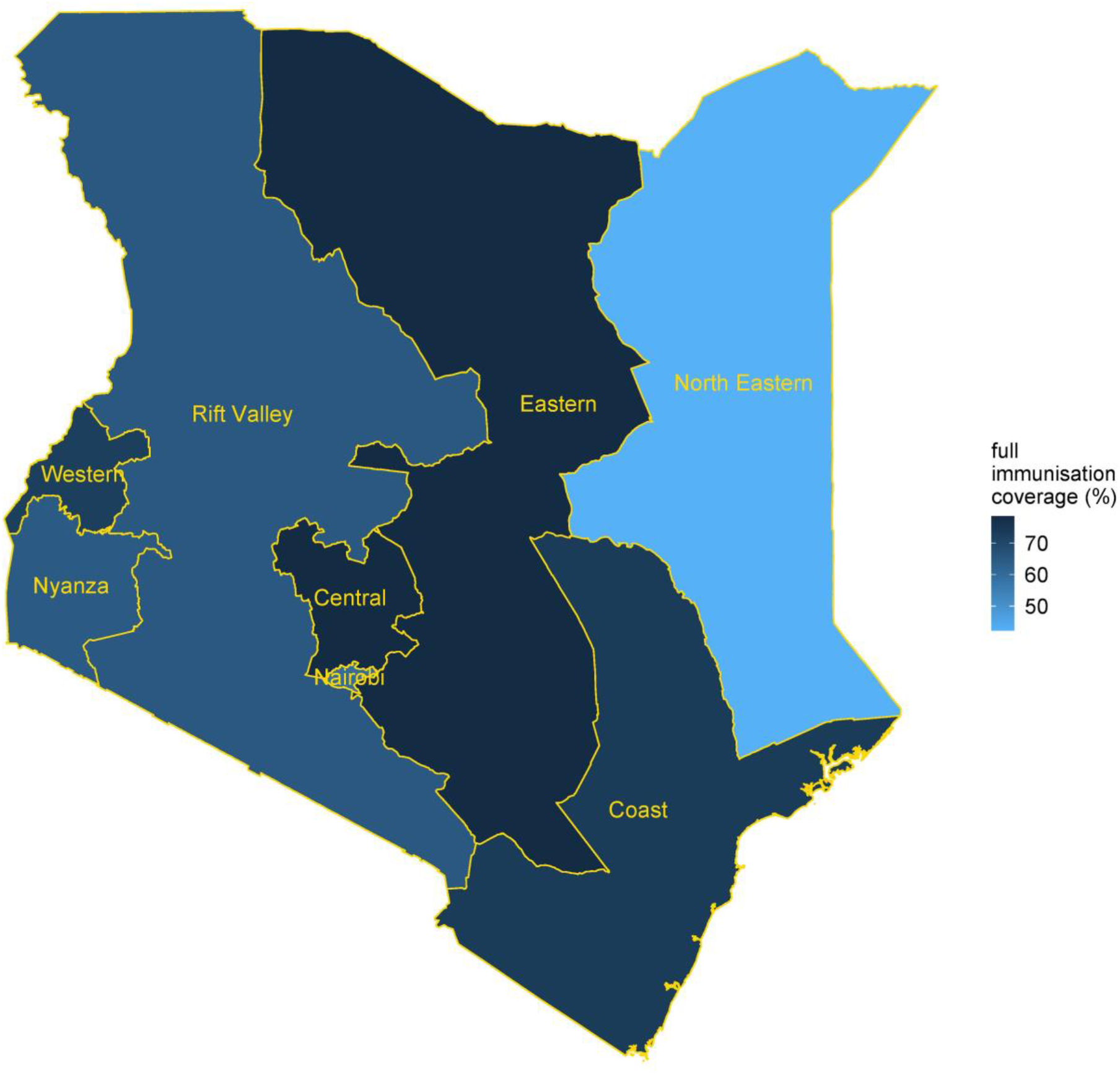
Full immunisation coverage in Kenya among children aged 12–23 months at the regional level. Full immunisation coverage (1-dose BCG, 3-dose DTP-HepB-Hib, 3-dose polio, 1-dose measles, and 3-dose pneumococcal vaccines) in Kenya among children aged 12–23 months in the eight regions of Kenya.

### Inequities in childhood immunisation coverage

Figure 2 presents the full immunisation coverage in Kenya among children aged 12–23 months disaggregated by socioeconomic (household wealth, religion, ethnicity), geographic (place of residence, region), maternal (maternal age at birth, maternal education, maternal marital status, maternal household head status), child (sex of child, birth order), and place of birth characteristics.

**Figure 2:**
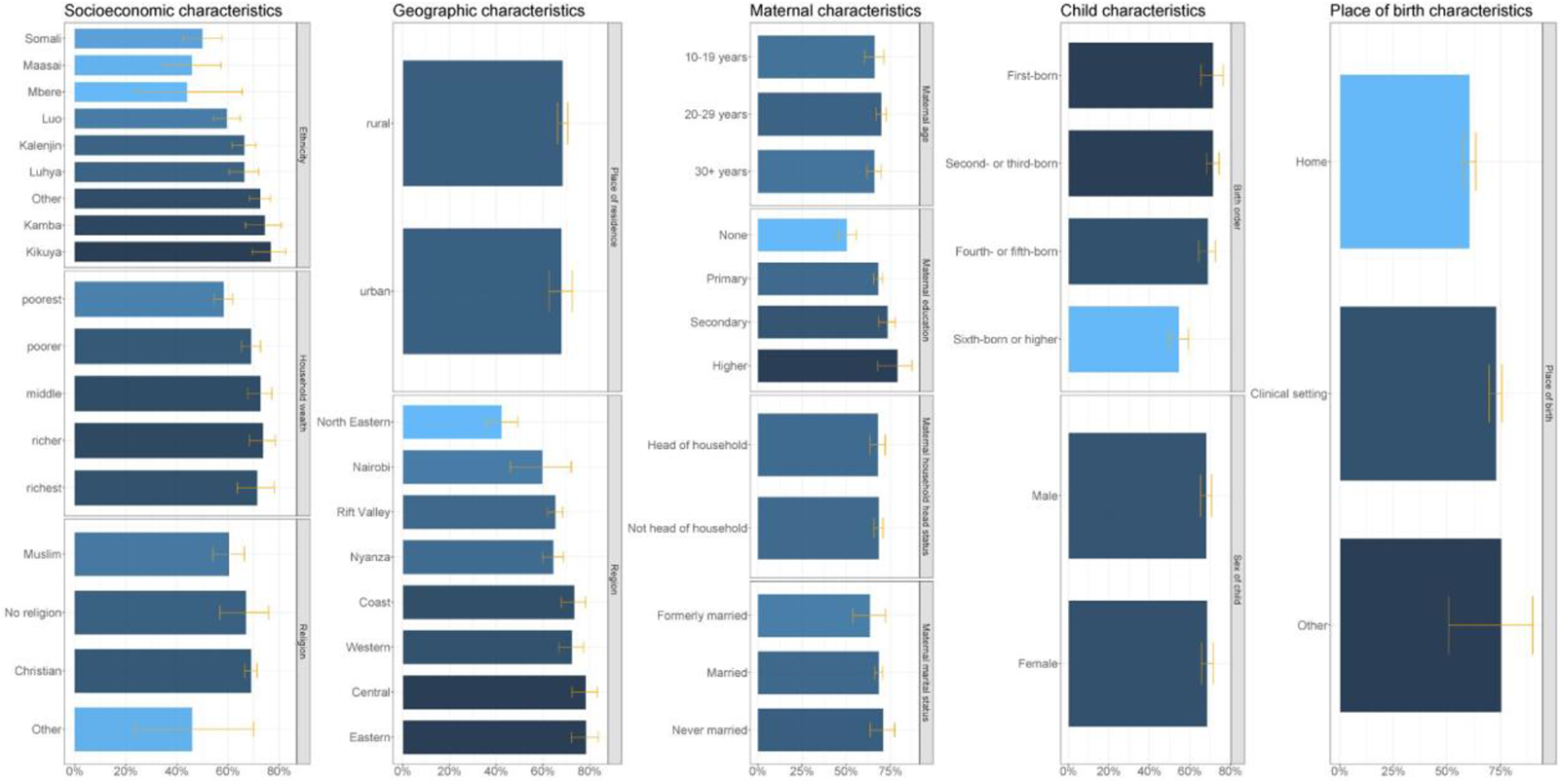
Full immunisation coverage in Kenya among children aged 12–23 months by socioeconomic, geographic, maternal, child and place of birth characteristics. Full immunisation coverage (1-dose BCG, 3-dose DTP-HepB-Hib, 3-dose polio, 1-dose measles, and 3-dose pneumococcal vaccines) in Kenya among children aged 12–23 months by socioeconomic (household wealth, religion, ethnicity), geographic (place of residence, province), maternal (maternal age at birth, maternal education, maternal marital status, maternal household head status), child (sex of child, birth order), and place of birth characteristics (x-axis refers to full immunisation coverage).

Ethnicity and religion was excluded from the multivariable logistic regression analysis due to collinearity with the region of residence – ethnic groups in Kenya tend to cluster in specific regions, and while the predominant religion in Kenya is Christianity, Muslims are predominantly based in the Coastal and North Eastern regions. Figure 3 present the adjusted odds ratios for full immunisation coverage in Kenya among children aged 12–23 months for socioeconomic (household wealth), geographic (place of residence, region), maternal (maternal age at birth, maternal education, maternal marital status, maternal household head status), child (sex of child, birth order), and place of birth characteristics.

**Figure 3:**
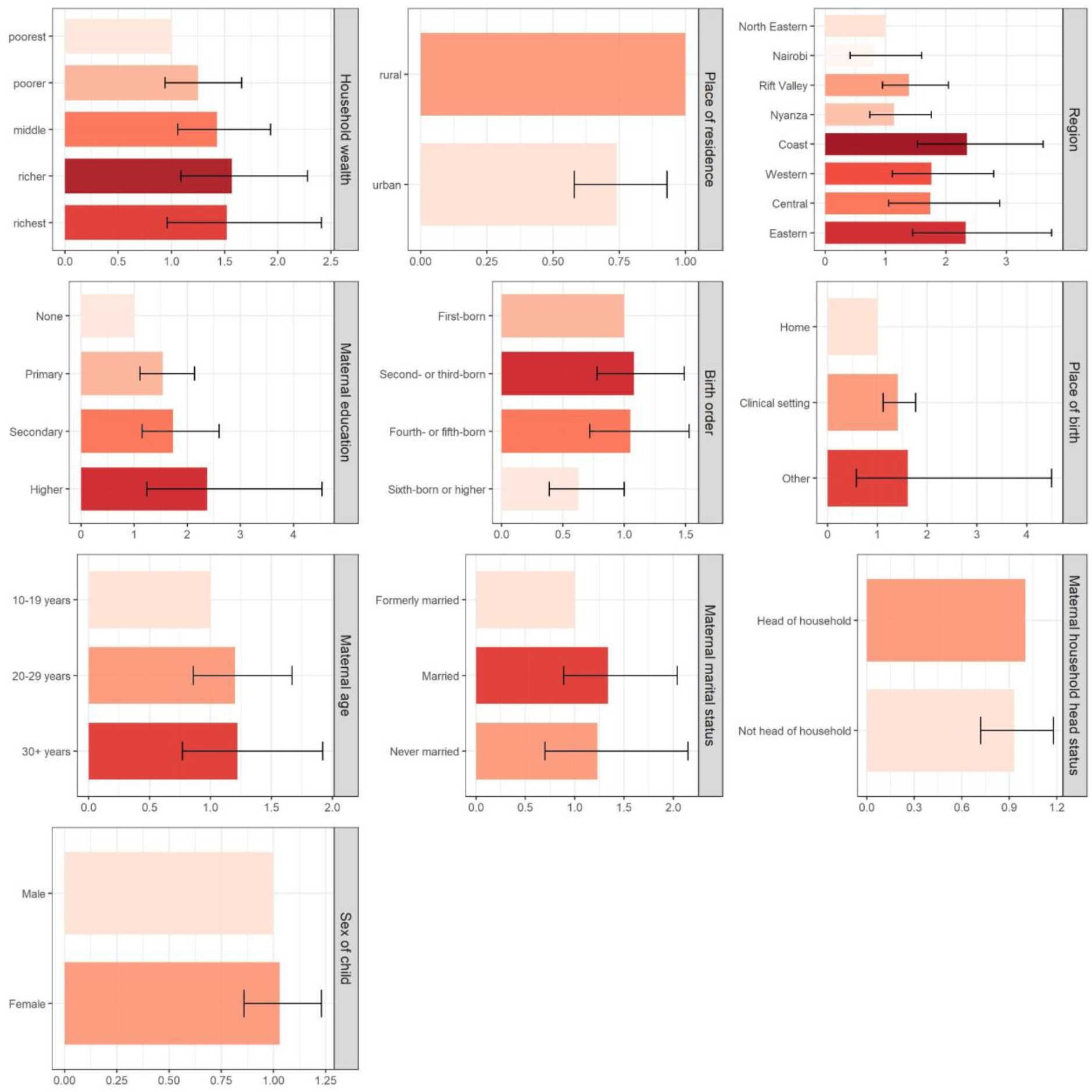
Inequities in full immunisation coverage in Kenya associated with socioeconomic, geographic, maternal, child, and place of birth characteristics. Inequities in full immunisation coverage (1-dose BCG, 3-dose DTP-HepB-Hib, 3-dose polio, 1-dose measles, and 3-dose pneumococcal vaccines) in Kenya among children aged 12–23 months associated with socioeconomic (household wealth), geographic (place of residence, province), maternal (maternal age at birth, maternal education, maternal marital status, maternal household head status), child (sex of child, birth order), and place of birth characteristics, based on multiple logistic regression estimates of adjusted odds ratios (x-axis refers to adjusted odds ratios).

Table 2 presents the crude odds ratios estimated by simple regression analysis and adjusted odds ratios estimated by multivariable regression analysis. After controlling for other background characteristics, we found strong evidence of association for maternal education and place of birth with full immunisation, moderate to very strong evidence of association for region and place of residence with full immunisation, moderate evidence of association for household wealth with full immunisation, and low evidence of association for birth order with full immunisation. Children of mothers with primary school education or higher have at least 54% higher odds of being fully immunised compared to children of mothers with no education. Children born in clinical settings have 41% higher odds of being fully immunised compared to children born in home settings. Children in the Coast, Western, Central, and Eastern regions had at least 74% higher odds of being fully immunised compared to children in the North Eastern region. Further, children in urban areas had 26% lower odds of full immunisation in comparison to children in rural areas. Children in the middle and richer wealth quintile households were 43–57% more likely to have full immunisation coverage in comparison to children in the poorest wealth quintile households. Children who were sixth born or higher had 37% lower odds of full immunisation compared to first-born children.

**Table 2:** Inequities in full immunisation coverage in Kenya associated with socioeconomic, geographic, maternal, child, and place of birth characteristics. Inequities in full immunisation coverage (1-dose BCG, 3-dose DTP-HepB-Hib, 3-dose polio, 1-dose measles, and 3-dose pneumococcal vaccines) in Kenya among children aged 12–23 months associated with socioeconomic (household wealth, religion, ethnicity), geographic (place of residence, province), maternal (maternal age at birth, maternal education, maternal marital status, maternal household head status), child (sex of child, birth order), and place of birth characteristics. Crude odds ratios were estimated by simple logistic regression and adjusted odds ratios were estimated by multivariate logistic regression. Ethnicity and religion were excluded from the multivariate logistic regression analysis due to collinearity with the region of residence.

| Characteristics | Children in each subgroup n (% of total) | Full immunisation coverage (%) | Crude odds ratio |  | Adjusted odds ratio |  |
| --- | --- | --- | --- | --- | --- | --- |
|  |  |  | OR and 95% CI | p-value | AOR and 95% CI | p-value |
| Household wealth (quintiles) |  |  |  |  |  |  |
| Poorest | 996 (25.3) | 58.3 (54.5, 61.9) | 1.00 | <0.001 | 1.00 | - |
| Poorer | 811 (20.6) | 69.0 (65.2, 72.7) | **1.60 (1.26, 2.03)** |  | 1.25 (0.94, 1.66) | 0.117 |
| Middle | 705 (17.9) | 72.7 (67.7, 77.1) | **1.90 (1.45, 2.50)** |  | **1.43 (1.06, 1.93)** | **0.020** |
| Richer | 684 (17.4) | 73.7 (68.2, 78.5) | **2.00 (1.47, 2.73)** |  | **1.57 (1.09, 2.28)** | **0.016** |
| Richest | 746 (18.9) | 71.3 (63.6, 78.0) | **1.78 (1.21, 2.62)** |  | 1.52 (0.96, 2.41) | 0.073 |
| Religion |  |  |  |  |  |  |
| Muslim | 336 (8.5) | 60.4 (54.1, 66.4) | 1.00 | 0.01 |  |  |
| No religion | 107 (2.7) | 67.0 (56.6, 75.9) | 1.33 (0.80, 2.20) |  |  |  |
| Christian | 3,488 (88.6) | 69.0 (66.6, 71.3) | **1.46 (1.12, 1.90)** |  |  |  |
| Other | 5 (0.1) | 46.0<br>(23.6, 70.0) | 0.56<br>(0.20, 1.58) |  |  |  |
| Ethnicity |  |  |  |  |  |  |
| Somali | 153<br>(3.9) | 50.0<br>(42.5, 57.5) | 1.00 | <0.001 |  |  |
| Maasai | 136<br>(3.5) | 45.9<br>(35.0, 57.2) | 0.85<br>(0.49, 1.46) |  |  |  |
| Mbere | 13<br>(0.3) | 43.9<br>(24.4, 65.5) | 0.78<br>(0.31, 1.99) |  |  |  |
| Luo | 497<br>(12.6) | 59.6<br>(54.2, 64.8) | **1.48**<br>**(1.00, 2.17)** |  |  |  |
| Kalenjin | 523<br>(13.3) | 66.4<br>(61.6, 71.0) | **1.98**<br>**(1.37, 2.86)** |  |  |  |
| Luhya | 604<br>(15.3) | 66.4<br>(60.4, 71.9) | **1.98**<br>**(1.33, 2.93)** |  |  |  |
| Kamba | 376<br>(9.5) | 74.4<br>(66.8, 80.9) | **2.91**<br>**(1.80, 4.71)** |  |  |  |
| Kikuya | 698<br>(17.7) | 76.7<br>(69.5, 82.6) | **3.29**<br>**(2.06, 5.25)** |  |  |  |
| Other | 943<br>(23.9) | 72.6<br>(68.3, 76.5) | **2.65**<br>**(1.83, 3.84)** |  |  |  |
| Place of residence |  |  |  |  |  |  |
| Rural | 2,591<br>(65.7) | 68.4<br>(66.2, 70.6) | 1.00 | 0.81 | 1.00 | - |
| Urban | 1,352<br>(34.3) | 67.8<br>(62.6, 72.5) | 0.97<br>(0.75, 1.25) |  | **0.74**<br>**(0.58, 0.93)** | **0.010** |
| Region |  |  |  |  |  |  |
| North Eastern | 129<br>(3.3) | 42.3<br>(35.6, 49.3) | 1.00 | <0.001 | 1 | - |
| Nairobi | 423<br>(10.7) | 59.7<br>(46.0, 72.1) | **2.02**<br>**(1.09, 3.77)** |  | 0.81<br>(0.41, 1.60) | 0.55 |
| Rift Valley | 1,128<br>(28.6) | 65.2<br>(61.8, 68.4) | **2.56**<br>**(1.86, 3.52)** |  | 1.39<br>(0.95, 2.04) | 0.09 |
| Nyanza | 574<br>(14.6) | 64.4<br>(59.8, 68.7) | **2.47**<br>**(1.75, 3.48)** |  | 1.14<br>(0.74, 1.76) | 0.54 |
| Coast | 407<br>(10.3) | 73.4<br>(67.8, 78.3) | **3.77**<br>**(2.55, 5.57)** |  | **2.35**<br>**(1.53, 3.61)** | **<0.001** |
| Western | 444<br>(11.3) | 72.4<br>(66.8, 77.4) | **3.59**<br>**(2.43, 5.30)** |  | **1.76**<br>**(1.11, 2.79)** | **0.016** |
| Central | 384<br>(9.7) | 78.3<br>(72.4, 83.3) | **4.94**<br>**(3.22, 7.57)** |  | **1.74**<br>**(1.05, 2.89)** | **0.031** |
| Eastern | 454<br>(11.5) | 78.4<br>(72.1, 83.6) | **4.95**<br>**(3.18, 7.70)** |  | **2.33**<br>**(1.45, 3.75)** | **0.001** |
| Maternal age at birth (years) |  |  |  |  |  |  |
| 10-19 | 522<br>(13.2) | 66.0<br>(60.2, 71.3) | 1.00 | 0.22 | 1.00 | - |
| 20-29 | 2,373<br>(60.2) | 69.8<br>(66.6, 72.8) | 1.19<br>(0.90, 1.58) |  | 1.20<br>(0.86, 1.67) | 0.27 |
| 30+ | 1,049<br>(26.6) | 65.8<br>(61.6, 69.7) | 0.99<br>(0.72, 1.36) |  | 1.22<br>(0.77, 1.92) | 0.40 |
| Maternal education |  |  |  |  |  |  |
| None | 459<br>(11.6) | 50.4<br>(45.1, 55.8) | 1.00 | <0.001 | 1.00 | - |
| Primary | 2,195<br>(55.7) | 68.1<br>(65.4, 70.6) | **2.09**<br>**(1.64, 2.68)** |  | **1.54**<br>**(1.11, 2.14)** | **0.009** |
| Secondary | 960<br>(24.4) | 73.3<br>(68.3, 77.8) | **2.70**<br>**(1.95, 3.74)** |  | **1.73**<br>**(1.15, 2.60)** | **0.008** |
| Higher | 329<br>(8.4) | 78.9<br>(67.6, 87.0) | **3.67**<br>**(1.97, 6.81)** |  | **2.37**<br>**(1.24, 4.54)** | **0.009** |
| Maternal marital status |  |  |  |  |  |  |
| Formerly married | 312<br>(7.9) | 63.4<br>(53.6, 72.2) | 1.00 | 0.44 | 1.00 | - |
| Married | 3,357<br>(85.1) | 68.4<br>(66.1, 70.7) | 1.25<br>(0.83, 1.88) |  | 1.34<br>(0.89, 2.04) | 0.16 |
| Never married | 274<br>(7.0) | 70.9<br>(63.4, 77.4) | 1.40<br>(0.83, 2.38) |  | 1.23<br>(0.70, 2.15) | 0.47 |
| Maternal household head status |  |  |  |  |  |  |
| Head of household | 716<br>(18.2) | 67.9<br>(63.3, 72.1) | 1.00 | 0.88 | 1.00 | - |
| Not head of household | 3,227<br>(81.8) | 68.3<br>(65.6, 70.8) | 1.02<br>(0.81, 1.29) |  | 0.93<br>(0.72, 1.18) | 0.54 |
| Sex of child |  |  |  |  |  |  |
| Male | 2,052<br>(52.0) | 67.9<br>(65.1, 70.6) | 1.00 | 0.74 | 1.00 | - |
| Female | 1,891<br>(48.0) | 68.5<br>(65.5, 71.4) | 1.03<br>(0.87, 1.21) |  | 1.03<br>(0.86, 1.23) | 0.763 |
| Birth order |  |  |  |  |  |  |
| First-born | 1,006<br>(25.5) | 71.2<br>(65.3, 76.4) | 1.00 | <0.001 | 1.00 | - |
| Second- or third-born | 1,589<br>(40.3) | 71.2<br>(68.0, 74.3) | 1.00<br>(0.74, 1.36) |  | 1.08<br>(0.78, 1.49) | 0.65 |
| Fourth- or fifth-born | 766<br>(19.4) | 68.6<br>(64.2, 72.6) | **0.88**<br>**(0.63, 1.24)** |  | 1.05<br>(0.72, 1.53) | 0.80 |
| Sixth-born or higher | 582<br>(14.8) | 54.4<br>(49.3, 59.3) | **0.48**<br>**(0.34, 0.68)** |  | **0.63**<br>**(0.39, 1.00)** | **0.052** |
| Place of birth |  |  |  |  |  |  |
| Home | 1,424<br>(36.2) | 60.3<br>(57.0, 63.4) | 1.00 | <0.001 | 1.00 | - |
| Clinical setting | 2,485<br>(63.1) | 72.7<br>(69.4, 75.7) | **1.75**<br>**(1.42, 2.17)** |  | **1.41**<br>**(1.12, 1.77)** | **0.003** |
| Other | 26<br>(0.7) | 75.2<br>(50.7, 90.0) | 2.00<br>(0.68, 5.92) |  | 1.61<br>(0.58, 4.50) | 0.36 |

We tested for interaction between maternal age at birth and maternal education, household wealth and maternal age at birth, household wealth and maternal education, and household wealth and place of residence. Full vaccination coverage was higher among children of mothers of age 20 years or older who have at least primary education in comparison to no education, among children living in poorest and richer households (that is first and fourth quintiles by household wealth) with mothers of at least primary education in comparison to no education, and among children living in the wealthier households (that is fourth and fifth quintiles by household wealth) of rural areas in comparison to urban areas.

## Discussion

We analysed full immunisation coverage among children aged 12-23 months in Kenya and estimated the inequities in full immunisation coverage associated with socioeconomic, geographic, maternal, child, and place of birth characteristics using data from the 2014 Kenya DHS survey. We found that inequities in full immunisation primarily affect children born into poorer households, born to mothers with no education and with many siblings/children in the same household, and in regions with limited health infrastructure. Belonging to richer versus poorer households, born to an educated versus uneducated mothers, and born in a clinical versus home setting are all associated with higher full immunisation coverage. Children in rural areas and the Rift Valley, Coast, Western, Central, and Eastern regions had higher full immunisation coverage while children who were sixth born or higher had lower full immunisation coverage.

Our findings complement the evidence from related cross-sectional studies [26]. Calhoun et al analysed data from Gem, Nyanza province, Kenya in 2003 and found that lower immunisation coverage among children aged 12-23 months was associated with lower maternal income, lower maternal education, and households with an absent parent [21]. Mutua et al analysed data from two slums of Nairobi in 2008 and found that incomplete childhood immunisation was associated with fewer household assets and expenditure, ethnicity, place of delivery, maternal education, and maternal age [22]. Masters et al used data from the 2014 Kenya DHS with a primary focus on the Somali ethnic community and found that childhood immunisation status was associated with wealth and place of birth [27]. Subaiya et al analysed the data from the national measles-rubella immunisation campaign for children aged 9 months to 14 years conducted in 2016 and found that immunisation coverage was strongly associated with children’s school attendance, maternal education, and household wealth [28]. Ifedayo et al analysed the Kilifi Health and Demographic Surveillance System and inferred that younger maternal age, more previous children, and delivery in hospital were associated with higher immunisation coverage and the strongest detrimental factor was the operational challenge of vaccine stock outs [29].

### Socioeconomic characteristics

Household wealth is a significant determinant of vaccine inequity with children in richer households more likely to be fully immunised than children in poorer households, even though vaccines are provided free-of-charge in public facilities in Kenya. This is consistent with previous studies which found that the poorest households face both financial and non-financial barriers to accessing immunization services [21,22,30]. The barriers include transportation cost to access the public facilities, childcare cost for other children, and the opportunity cost of taking time off work. New approaches for delivering immunization services to reduce the travel time, such as constructing new health facilities in underserved areas or introducing community health worker models that operate on a localized level will facilitate improved access to immunisation services [31].

### Geographic characteristics

Children living more than two hours away from health facilities providing immunisation are less likely to be fully immunised and receive DPT3 after controlling for household wealth, mother’s highest education level, parity and urban/rural residence [32]. Of Kenya’s 47 counties, 29 counties do not meet the national policy target of 90% of the population living within one hour at walking speed of a health facility offering immunisation services [32,33]. But, we inferred that children living in urban areas were less likely to be immunised than children in rural areas. Similar associations have been found in previous studies, with residents of urban slums, who are typically much poorer than other urban residents, often driving part of the inequities [21,34]. Residents in urban slums have limited access to employment, water and sanitation, are inadequately served with basic public services such as immunisation and education, and have the worst health and socio-economic outcomes among all social groups in Kenya [35]. Urban slums also tend to have higher rates of population growth compared to non-slum urban areas, which further exacerbates the relative inequities in childhood immunisation coverage.

As inferred by the multicollinearity between the defunct provinces and ethnicity, the regional differences in immunisation coverage are closely linked to the distribution of different ethnic groups across Kenya. The Central province has a large proportion of people of Kikuyu ethnicity and children have higher rates of immunisation coverage [36]. The North Eastern province borders Somalia and is home to millions of people of Somali ethnicity who have the lowest immunisation coverage of any ethnic group in Kenya [27]. Migrants from Somalia – whether refugees, asylum seekers, or economic migrants – find it harder and are typically more reluctant to access public services such as immunization services due to discrimination and unfamiliarity with the system [37]. The North Eastern province is also home to the Dadaab refugee camp which houses more than 200,000 Somali refugees and is one of the largest refugee camps in the world [38]. Low immunisation rates in the refugee camp has led to prior outbreaks of vaccine-preventable diseases [39].

In 2010, Kenya adopted a new constitution that devolved administrative powers, including the responsibility for health and health care, to 47 county governments which are one administrative level below the now-defunct provincial system. This decentralization of power presents a critical opportunity for county governments in provinces with low immunisation coverage to reprioritize and redistribute funds towards improving the availability and accessibility of immunization services, particularly to marginalised populations.

### Maternal characteristics

Maternal education is a strong predictor of full immunisation coverage and is a consistent finding across related studies in Kenya and other countries [40,41]. Mothers with at least some education are more aware of the importance of childhood immunisation, either through education or by being exposed to school-based immunization programs themselves [30].

### Child characteristics

Being born into a family with few other children was associated with full immunisation, and has also been inferred by related studies in Kenya [42,43]. Parents with fewer children have more time to care for each child and are less likely to need to organize childcare for their other children while they travel to a health facility for immunization.

### Place of birth characteristics

Children born in clinical settings and health facilities in Kenya are more likely to be fully immunised than the children born in home settings, as also observed in related studies [27,44]. Health workers at clinical facilities are more likely to vaccinate the children with the birth dose of BCG and inform mothers on the recommended immunisation schedule in comparison to traditional birth attendants who support home-deliveries in Kenya [27,45]. Also, mothers giving birth in clinical settings practice relatively higher health-seeking behaviours, including immunisation for themselves and their children [42].

### Missed opportunities for vaccination

Coverage for vaccines in the first few months after birth, including BCG at birth and three doses of the pentavalent vaccine, have higher coverage than those administered towards the end of the first year such as the first dose of measles containing vaccine in Kenya. Thereby, concerted effort is required to keep mothers engaged with health facilities and immunisation services after the first few weeks post-birth. Missed opportunities for vaccination occur when children have contact with health services either directly, or indirectly through attending with family, but do not receive vaccine doses for which they are eligible [46]. They are attributed in part due to knowledge gaps in the routine immunisation schedule and issues in vaccine supply and vaccine-related equipment such as syringes and vaccination record books [47].

### Limitations

We did not include the rotavirus vaccine or the second-dose of measles vaccine in our analysis, as both were introduced after the 2014 Kenya DHS survey - in 2014 and 2015, respectively [48]. We are unable to infer temporal inferences and causal-effect relationships due to the cross-sectional study design. Our study is also subject to similar biases associated with DHS surveys, such as measurement bias, recall bias, and social desirability bias which tend to overestimate immunisation coverage. We did not disaggregate vaccination status data by vaccination card versus maternal recall in our analysis. For 25% of children in this study, data on vaccination status was based on maternal recall as written vaccination records were unavailable; the likelihood of missing written records has been found to be unevenly distributed across sub-groups whose characteristics may also be associated with vaccination coverage [49]. Vaccination coverage data collected through household surveys such as DHS do not always align with similar data collected through serological surveys [49].

There were similarities and differences in immunisation coverage estimates for Kenya among children aged 12-23 months based on a nationally representative sample of 3,943 children used in this study and 3,777 children in the Kenya DHS 2014 report [18], as shown in the appendix Table A1. The immunisation coverage estimates were similar for most vaccines – BCG scheduled at birth; 1^st^, 2^nd^, and 3^rd^ doses of DTP-HepB-Hib scheduled at 6, 10, 14 weeks; 1^st^, and 2^nd^ doses of polio scheduled at 6 and 10 weeks; 1^st^, 2^nd^, and 3^rd^ doses of pneumococcal scheduled at 6, 10, 14 weeks, and measles 1^st^ dose scheduled at 9 months of age. The coverage estimates were different for the 3^rd^ dose of polio and full immunisation. The reason for the differences in polio coverage, which in turn impacts the full immunisation coverage, was that in the Kenya DHS 2014 report – for children whose mothers reported that they had received three doses of DPT-HepB-Hib and polio 0, polio 1, and polio 2, it was assumed that polio 0 was in fact polio 1, polio 1 was polio 2, and polio 2 was polio 3, while we did not make this assumption in our analysis.

### Future directions

We need implementation research and evidence on interventions that would reduce inequities in childhood immunisation in Kenya and to inform the redistribution of healthcare resources to protect all children with full immunisation, regardless of location, age, socioeconomic status or gender-related barriers [10,50]. Specifically, qualitative research to infer the barriers faced by families of under-immunised children to accessing vaccination would be valuable to inform and adapt immunisation services to overcome these barriers. Designing cost-effective solutions to reduce inequities in immunisation coverage between different regions, between rural and urban areas, and between richer, more highly educated mothers and poorer, less educated mothers would be beneficial. Also, Kenya DHS 2021 is ongoing, and sampling is conducted in two stages to achieve representativeness at the national, regional, and county levels at a more granular level in comparison to Kenya DHS 2014. Disaggregated data at the county level will be valuable for the 47 administrative counties that form the core decision-making structures for health since the devolution of power in 2010.

### COVID-19 pandemic impact on disruption of immunisation services in Kenya

The COVID-19 pandemic has disrupted routine childhood immunisation and led to the suspension of supplementary immunisation activities in many countries including Kenya [51–53]. Immunisation services have been disrupted in both fixed post and outreach immunisation activities due to health workers being redeployed to the COVID-19 response and parents being unable or unwilling to bring their children into health facilities because of restrictions on movements, economic hardships, or the fear of contracting SARS-CoV-2 while attending health facilities, among other reasons [51]. These disruptions to immunisation services are likely to expand the equity gap and this should receive attention as part of efforts to restore health services and provide catch-up vaccination.

### Conclusions

The inequities in full immunisation coverage are primarily affecting children born into poorer households, to mothers with no education and with many other children, and in provinces with limited health infrastructure. These under-immunised children, who are already at a socioeconomic disadvantage in early life, are more susceptible to infectious diseases which worsens their early childhood development with potential lifelong sequelae or death. Further, while the COVID-19 pandemic has disrupted routine and campaign immunisation services in 2020, it also presents an opportunity to tackle the identified inequalities as immunisation services are restored to capacity.

## Data Availability

The analysis code is publicly accessible at https://github.com/vaccine-impact/vaccine_equity_kenya and the 2014 Kenya DHS data set is accessible upon registration on the DHS website at https://www.dhsprogram.com/methodology/survey/survey-display-451.cfm.

## Declarations

### Ethics approval and consent to participate

This study was approved by the ethics committee (Ref 19139) of the London School of Hygiene & Tropical Medicine. The DHS Program obtained informed consent for all DHS survey respondents or, if subjects are under 18, from a parent and/or legal guardian. All methods were carried out in accordance with relevant guidelines and regulations.

### Consent for publication

Not applicable.

### Availability of data and materials

The analysis code is publicly accessible at https://github.com/vaccine-impact/vaccine_equity_kenya and the 2014 Kenya DHS data set is accessible upon registration on the DHS website at https://www.dhsprogram.com/methodology/survey/survey-display-451.cfm. To download DHS datasets, researchers must register as a DHS data user at https://dhsprogram.com/data/new-user-registration.cfm.

## Authors’ contributions

SA and KA conceptualised the study and SA undertook the analysis and wrote the initial draft. KA and IMOA contributed to the analysis, interpretation of results, and the final drafting of the manuscript.

## Acknowledgments

We thank the Demographic and Health Surveys (DHS) Program for access to the 2014 Kenya DHS data set.

### Appendix

**Table A1:**
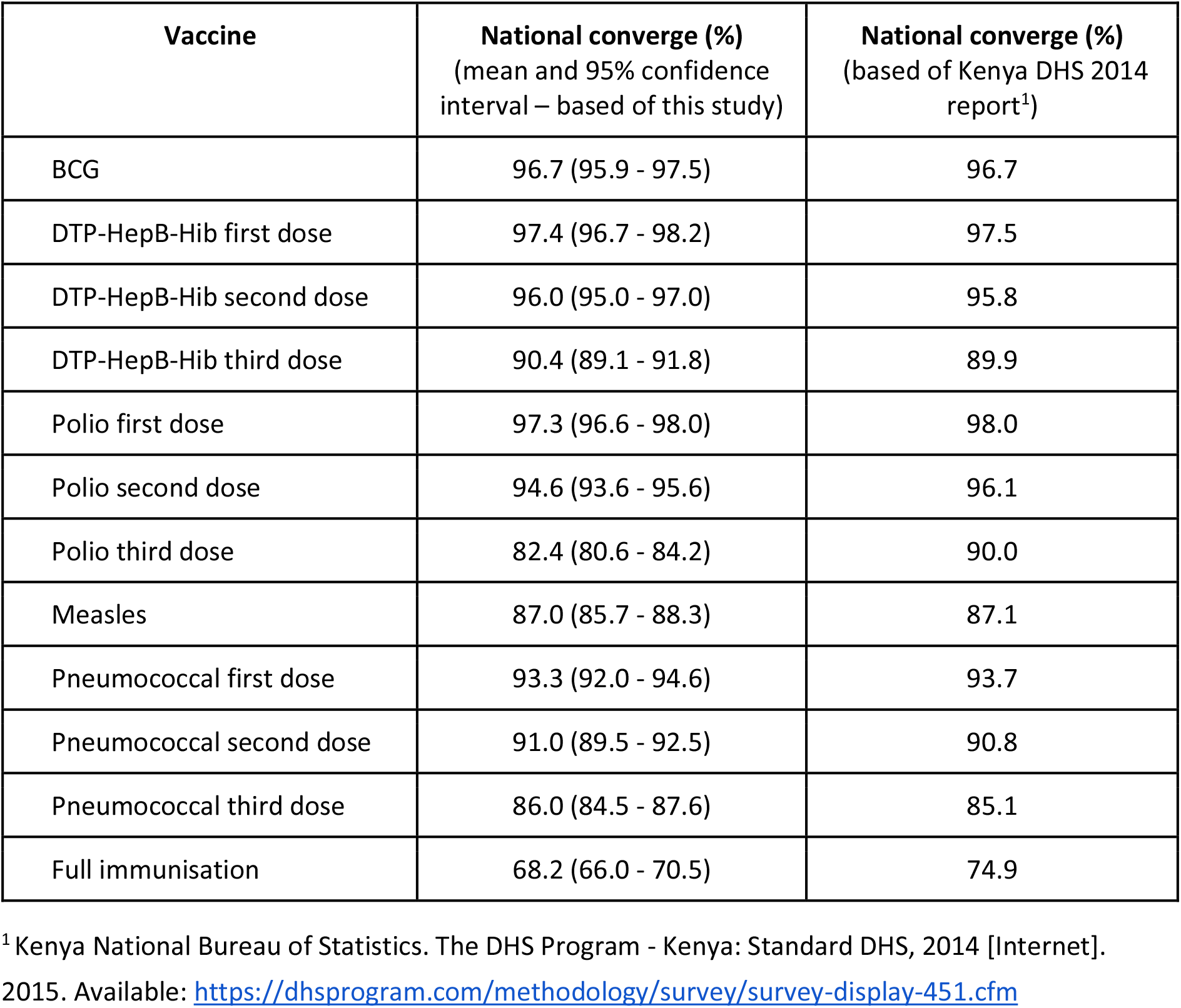
Similarities and differences in immunisation coverage estimates for Kenya. Immunisation coverage in Kenya among children aged 12-23 months based on a nationally representative sample of 3,943 children used in this study and 3,777 children in the Kenya DHS 2014 report^1^. Full immunisation includes 1-dose BCG, 3-dose DTP-HepB-Hib, 3-dose polio, 1-dose measles, and 3-dose pneumococcal vaccines.

| **Vaccine** | **National converge (%)**<br>(mean and 95% confidence interval – based of this study) | **National converge (%)**<br>(based of Kenya DHS 2014 report<sup>1</sup>) |
| --- | --- | --- |
| BCG | 96.7 (95.9 - 97.5) | 96.7 |
| DTP-HepB-Hib first dose | 97.4 (96.7 - 98.2) | 97.5 |
| DTP-HepB-Hib second dose | 96.0 (95.0 - 97.0) | 95.8 |
| DTP-HepB-Hib third dose | 90.4 (89.1 - 91.8) | 89.9 |
| Polio first dose | 97.3 (96.6 - 98.0) | 98.0 |
| Polio second dose | 94.6 (93.6 - 95.6) | 96.1 |
| Polio third dose | 82.4 (80.6 - 84.2) | 90.0 |
| Measles | 87.0 (85.7 - 88.3) | 87.1 |
| Pneumococcal first dose | 93.3 (92.0 - 94.6) | 93.7 |
| Pneumococcal second dose | 91.0 (89.5 - 92.5) | 90.8 |
| Pneumococcal third dose | 86.0 (84.5 - 87.6) | 85.1 |
| Full immunisation | 68.2 (66.0 - 70.5) | 74.9 |

<sup>1</sup> Kenya National Bureau of Statistics. The DHS Program - Kenya: Standard DHS, 2014 [Internet]. 2015. Available: https://dhsprogram.com/methodology/survey/survey-display-451.cfm

